# Consent-driven, semi-automated data collection during birth and newborn resuscitation: Insights from the NewbornTime study

**DOI:** 10.1101/2025.01.15.24319287

**Authors:** Sara Brunner, Anders Johannessen, Jorge García-Torres, Ferhat Özgur Catak, Øyvind Meinich-Bache, Siren Rettedal, Kjersti Engan

**Author notes:** Corresponding author: (SB).

## Abstract

Accurate observations at birth and during newborn resuscitation are fundamental for quality improvement initiatives and research. However, manual data collection methods often lack consistency and objectivity, are not scalable, and may raise privacy concerns. The NewbornTime project aims to develop an AI system that generates accurate timelines from birth and newborn resuscitation events by automated video recording and processing, providing a source of objective and consistent data. This work aims to describe the implementation of the data collection system that is necessary to support the project’s purpose.

Videos were recorded using thermal cameras in labor rooms and thermal and visual light cameras in resuscitation rooms. Consent from mothers were obtained before birth, and healthcare providers were given the option to delete videos by opting out. The video collection process was designed to minimize interference with ongoing treatment and not impose unnecessary burden on healthcare providers. Videos have been collected at Stavanger University Hospital since November 2021. By July 31^st^ 2024, 645 thermal videos of birth and 186 visual light videos of resuscitation have been collected. Data collection and development and implementation of AI systems is still ongoing.

The utilization of automated data collection and AI video processing around birth may allow for consistent and enhanced documentation, quality improvement initiatives, and research opportunities on sequence, timing and duration of treatment activities during acute events, with less efforts needed for capturing data and improved privacy for participants.

**Author summary:** In our study, we address the need for accurate and consistent data collection during birth and newborn resuscitation. Traditional manual data collection methods often fall short in terms of objectivity, consistency, scalability, and privacy. To overcome these challenges, we have developed a consent-driven, semi-automated data collection system with the aim of developing AI-based systems that uses video recordings to create precise timelines of events occurring at birth and during newborn resuscitation. This method leverages thermal and visual light cameras to capture footage, ensuring minimal interference with clinical practices and respecting the privacy of participants. This work describes the detailed setup of the data collection and sets it in perspective to existing solutions used in clinical practice and research. Since November 2021, we have collected data at Stavanger University Hospital, including 645 thermal videos of births and 186 visual light videos of resuscitation by the end of July 2024. This data will support the future development of AI algorithms aimed at enhancing the objectivity and consistency of data collection. Ultimately objective data can be used for individual learning or, on a big scale, understanding adherence to guidelines and effect of treatment, when combined with outcome data.

## Introduction

Data collection during birth and newborn resuscitation presents challenges. Manual observations in real time and annotation of video observations may have issues with objectivity, accuracy, privacy and scalability [1–4]. Accurate and high-quality data is needed for documentation, research, quality improvement initiatives and for learning purposes. Recently, Bettinger et al. [5] addressed the importance of making “every birth a learning event” to improve the competence of healthcare providers, as they seldom encounter newborn resuscitation in their clinical practice.

The NewbornTime project [6], registered in ISRCTN Registry [7], is a collaborative project between University of Stavanger (UiS), Stavanger University Hospital (SUH), bitUnitor AS and Laerdal Medical AS that aims to improve data collection from birth and during newborn resuscitation and to increase the value of collected data. The overall objective is to develop an AI system that uses video input to automatically generate timelines of events and activities, starting with time of birth (ToB) and continues throughout newborn stabilization and resuscitation.

In this work, we describe a semi-automated data collection system based on digitally stored informed consent of participants (mothers) and an opt-out option for healthcare providers. Our solution does not interfere with clinical practice and adds minimal burden to healthcare providers.

## Method

All videos are collected at the labor ward at SUH. The hospital has fourteen labor rooms at the labor ward and one operation theater used for C-sections situated nearby. There are four additional labor rooms for low-risk labors, that were not included for video collection in this project. Approximately 4000 newborns are born each year at SUH, about 10% need help to start breathing, and 3-4% receive positive pressure ventilation (PPV) [8]. Newborns receiving treatment exceeding drying, are placed on a resuscitation station in a treatment room close to the labor rooms. Newborns born by C-section are treated on a resuscitation station in the anteroom in conjunction with the operation theater.

Development of AI algorithms for activity recognition in resuscitation videos and detection of birth from thermal videos is done by researchers at UiS. This work is ongoing, but some results have been published [9–11]. UiS is also developing methods for exploring activity timelines [12].

### Ethics Statement

The study has been approved by the Regional Ethical Committee, region west, Norway (REK-Vest), REK number: 222455. Written consent has been obtained from all participating mothers. Healthcare providers where not defined as study participants and no consent was obtained. The study is registered in the ISRCTN registry: ISRCTN12236970.

### Participants

All women who give birth at SUH during the study period and have consented are eligible for participation. Pregnant women are informed about and invited to participate in the study during their antenatal visits around pregnancy weeks 12 and 20. Furthermore, if the midwives have the capacity, pregnant women will be asked to participate upon arrival at the labor ward.

### Digital, self-serviced consent

The Digital Consent Platform was developed by bitUnitor AS, Norway, based on blockchain using Hyperledger technology hosted on a cloud service provider. It ensures the secure storage and encryption of personal information for participating mothers, and it creates study IDs if consent and data is present. To ensure immutability, the consent status is stored within a blockchain. The digital consent platform provides a protected API that enables the retrieval of consent status and study ID by authorized applications. The cloud service provides robust security measures for APIs, including centralized access control, encryption, IP address restrictions, and continuous threat monitoring. Data is encrypted both in transit and at rest, ensuring protection against interception and unauthorized access. Additionally, IP address restrictions and virtual network integration, narrowing access to trusted sources only and minimizing potential attack vectors.

During the study period, an information video was shown continuously in the two waiting areas for antenatal care at SUH, where also paper brochures about the study were available. Most days a research assistant or a nurse assistant would be available to explain pregnant women about the study. All informed the pregnant women about the possibility to consent digitally, by scanning a QR code and filling in an online form. Alternatively, they could also sign the paper consent in the brochure, which would later be entered into the Digital Consent Platform by a research assistant.

### Thermal (infrared) cameras

The study utilizes 12 thermal camera sensors Mx-O-SMA-TPR079 connected to Mx-S16B camera modules (both Mobotix AG, Germany) to capture video from the labor and treatment rooms. The sensors provide a frame rate of 8.33 frames per second, with an image size of 336 × 252 pixels, and have a field of view measuring 45° × 35°. The sensors are not equipped with a microphone.

The thermal sensors are linked to camera modules via USB cables, and a subsequent wired connection connects the thermal cameras to a secure local network. A Linux server within the same network interacts with the thermal cameras over HTTP using the MXCameraSystem SDK 1.0.2 (Mobotix AG, Germany) software.

An event is set up on the thermal cameras that triggers once every 5 seconds if the thermal sensor detects at least one pixel above 30° C. This event is used to start data collection for 20 seconds. To allow for continuous data collection the 20 seconds timer is restarted every time the temperature event is triggered.

Thermal raw data is stored on a RAID (Redundant Array of Independent Disks) storage together with metadata detailing information about the camera, including the room where the camera is located, the location of the sensor relative to the bed (“head” / “side”), and the recording start time. This information is used when processing the data where the raw data is divided into one file per frame and uploaded to the permanent cloud infrastructure. This file structure allows for easy conversion to video.

Thermal cameras offer several advantages for detecting the ToB in labor rooms. These cameras sense thermal radiation in the infrared spectrum and capture significantly fewer details compared to visible light cameras, thereby enhancing privacy for both healthcare providers and women in labor. At the moment of birth, the newborn will be slightly warmer than the normal skin temperature of other people in the room, which can be exploited for detection of newborn ToB [10]. The detection process involves training Artificial Intelligence (AI) models on thermal video data, either frame-by-frame or by incorporating spatiotemporal information. How thermal imaging is working and what is affecting its accuracy in this setup has been described by García-Torres et al [13].

Figure 1 shows the sensor placement in the labor room and operational theater, respectively and two examples of a thermal image from actual sensors at the ToB.

**Figure 1.**
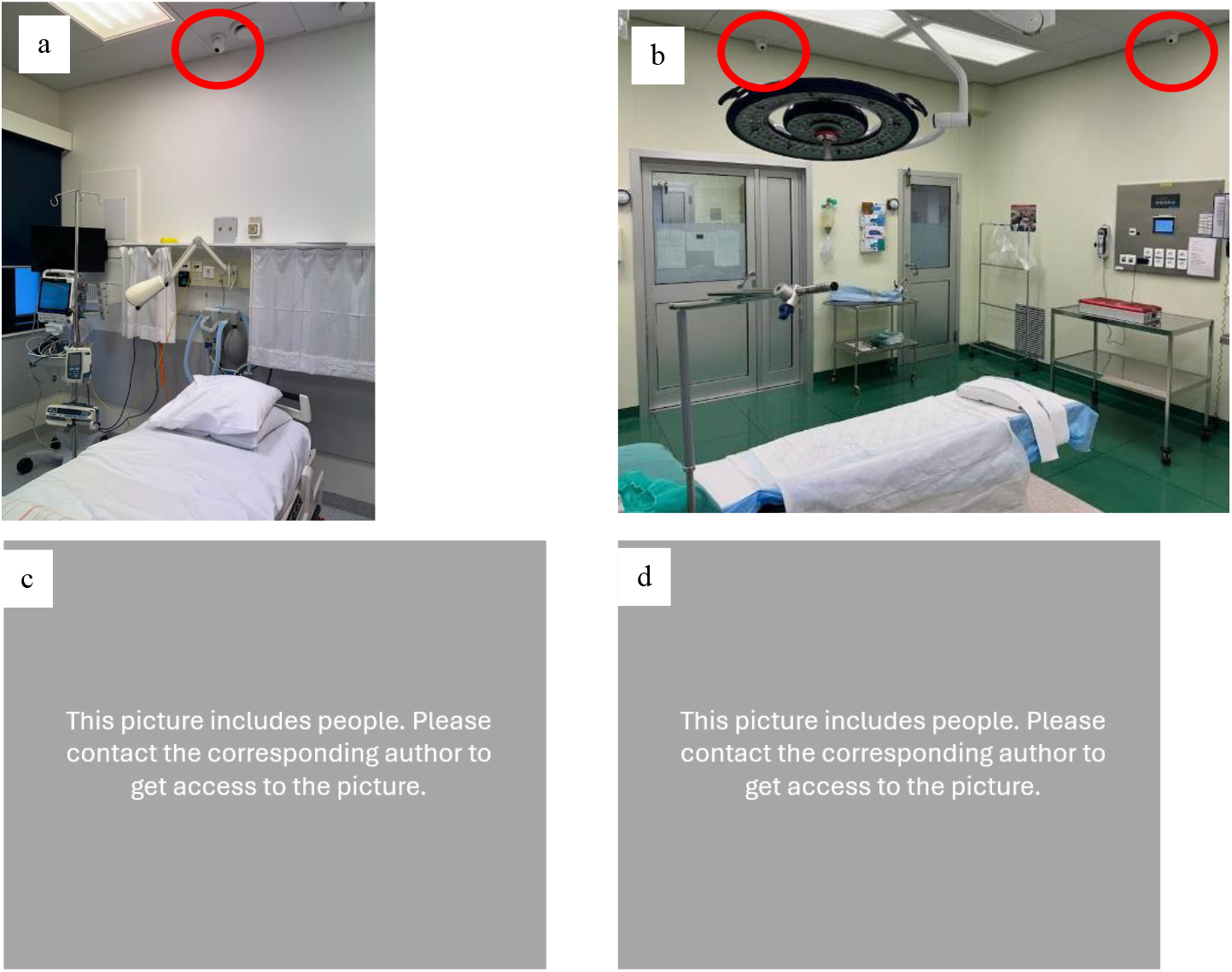
Top: Position of the thermal sensors (marked with a red circle) in a) a labor room, b) the operation theater for C-sections. Bottom: Thermal image at birth. c) from a vaginal delivery in the labor room, d) from a C-section delivery in the operating theater (distance between thermal sensor and operating table is bigger than in the labor rooms). The newborn is marked with a black circle.

Thermal cameras are also employed as overview cameras in the resuscitation rooms, see Figure 2 c). The choice of thermal cameras is again made to ensure the privacy of healthcare providers. These videos will document the number of healthcare providers present during newborn resuscitation.

**Figure 2.**
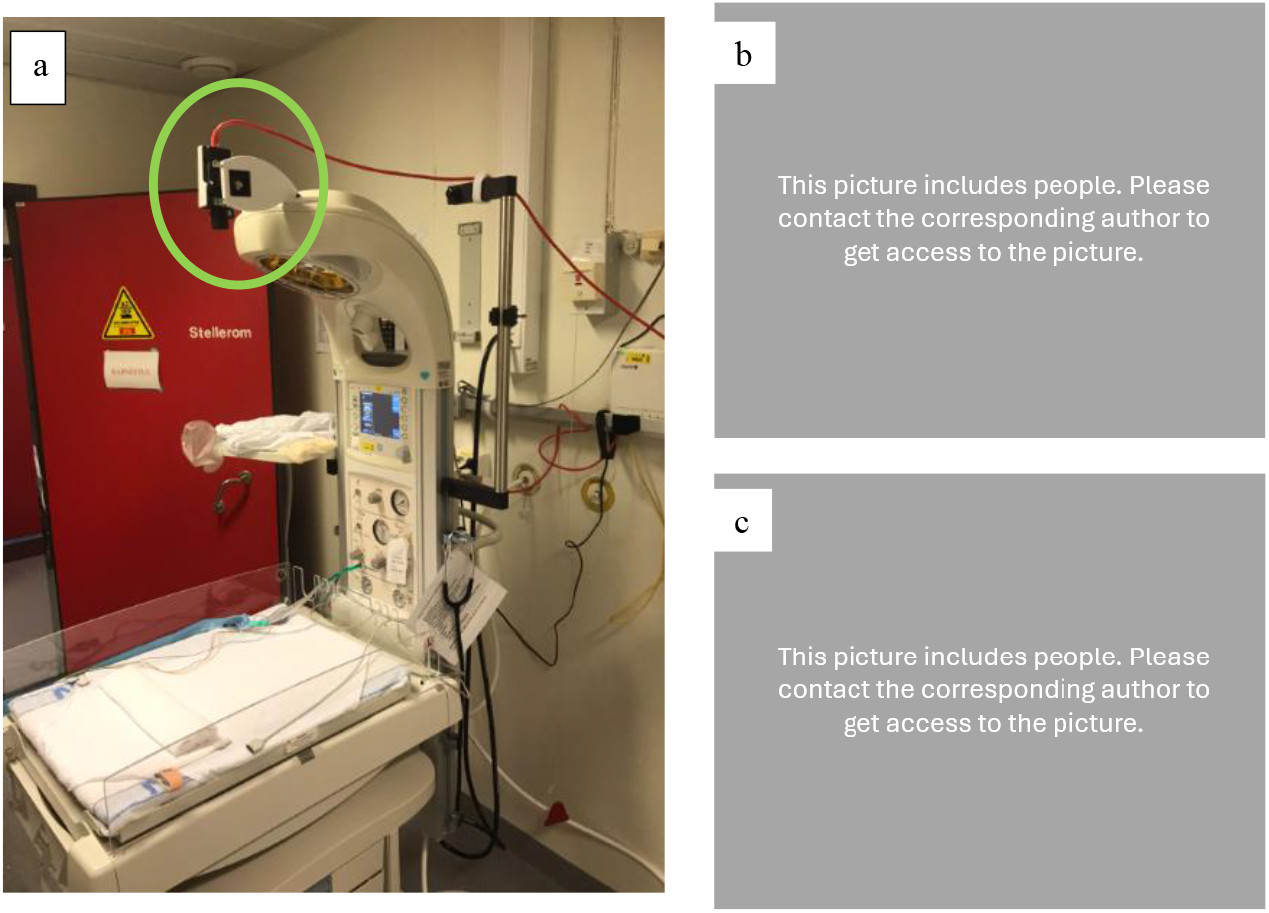
The visual light camera (green circle) is mounted directly above the treatment area (a). Treatment of newborns are captured well with the limited field of view (b) and the thermal camera captures what is happening around the treatment area (c).

### Visual light cameras

The study utilizes three Axis M1134 Network Cameras (Axis Communications AB, Sweden). These cameras operate at a frame rate of 25 frames per second and have a resolution of 1024 × 768 pixels, providing a field of view measuring 90° × 49°.

The camera’s field of view is adjusted to encompass the smallest possible area of the resuscitation station, ensuring that all treatment actions are captured, while respecting privacy of clinical staff. Camera placement of the visual light camera and two image examples from treatments are shown in Figure 2.

The videos captured by these cameras are directly streamed to a RAID storage. Recording begins automatically when motion is detected within the camera’s field of view and continues for 5 minutes after the last motion detection. The recording also includes the 60 seconds video footage right before the first motion trigger. The sound is removed as part of the post-processing steps on the local server at SUH

Visual light videos are used to train AI models in identifying ongoing Newborn Resuscitation Algorithm Activities such as stimulation and PPV [9,11].

### Camera placement

#### Before May 2023

Four labor rooms were equipped with two thermal sensor modules each. One sensor was positioned on the ceiling directly above and behind the laboring woman’s head (“head-view”), while the other was placed on the right side of the woman in labor (“side-view”). Furthermore, two thermal sensors were installed on the ceiling in the operating theater for C-sections to obtain a “head-view” and “side-view” with minimal obstructions in the field of view.

All three, currently utilized resuscitation stations, were equipped with top-down visual light cameras, enabling recording of the surface area of the infant bed. This configuration ensures an unimpeded view of the newborn while minimizing the capture of personal characteristics of the clinicians, such as faces or ID cards. Furthermore, a thermal sensor has been strategically positioned on the ceilings of the treatment room and the anteroom to monitor the ongoing activities within these areas.

#### Since May 2023

Collection of thermal videos was extended to 8 labor rooms in May 2023. The “side-view” thermal sensor was removed from all labor rooms and installed as “head-view” sensor in four labor rooms that were previously not equipped with thermal sensors.

During the analysis of the thermal videos collected in the initial study period, it was noted that the moment of birth was visible in most cases from the “head view” data. In instances where it was not visible from the “head-view,” it was neither visible from the “side-view.” Based on these findings, the decision was made to relocate the “side-view” sensor to labor rooms that were not yet equipped with any sensor, to increase the number of recorded births. No modifications have been made to any visual light camera or the thermal sensors in the operating theater, treatment room and anteroom.

#### Liveborn Observation App

The Liveborn Observation App [14] (Figure 3), developed by Laerdal Medical AS, Norway, is installed on three Android tablets (Samsung Galaxy Tab A7 Lite) in the labor ward. Once delivery is expected, a midwife assistant brings a tablet to the labor room. In real-time, the midwife assistant annotates the time of birth and the time of cord clamp by pressing the respective on-screen buttons in the app.

**Figure 3.**
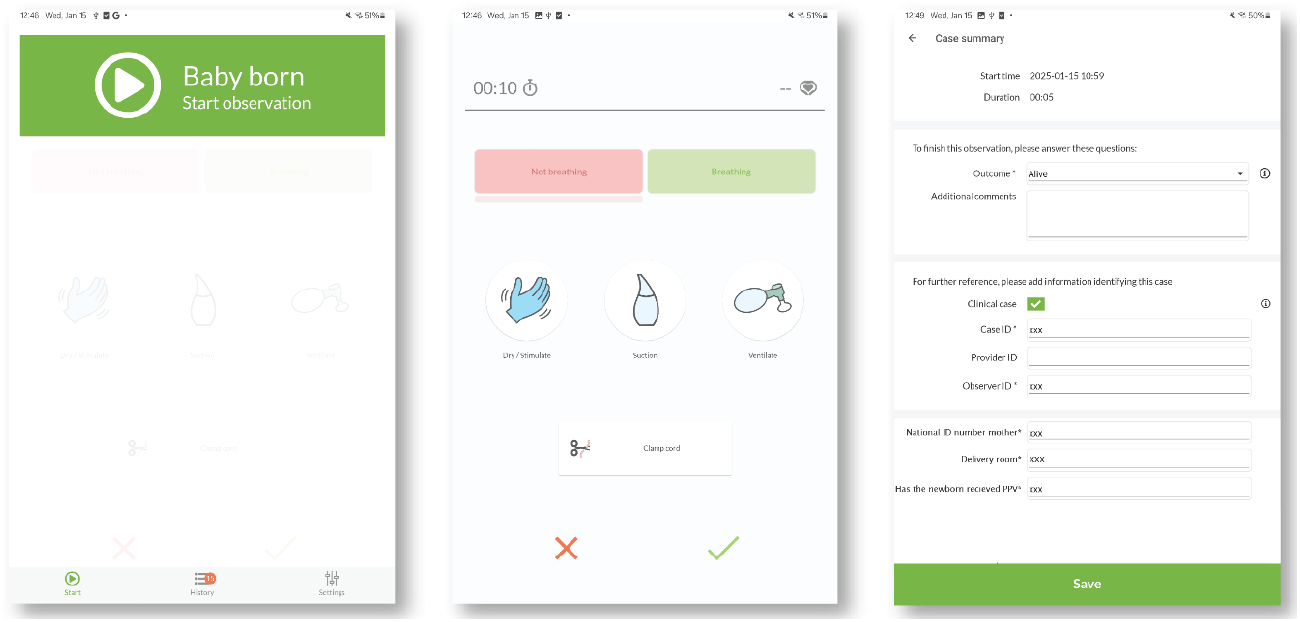
Screenshots of the Liveborn Observation app. Left: start screen where “Baby born” button is pressed to record the time of birth. Middle: Event registration screen with “Clamp cord” button, Right: Example of summary screen, where research assistant will add mothers personal ID and labor room number.

A research assistant adds additional case information to the app, such as the labor room and the personal ID number of the mother, typically within 24 hours.

Once the personal ID number of the mother is entered and confirmed, the app checks consent status in the Digital Consent Platform. If maternal consent is active, the app receives a unique study ID and adds it to the dataset. All collected app data is sent to the Liveborn Web application (Laerdal Medical AS, Stavanger). The personal ID number is deleted immediately after the consent check, independently of the received consent status and is never uploaded to the Liveborn web application.

#### Employees opt-out

Employees are not defined as study objects but have the option to request deletion of videos in which they have participated, within 48 hours after the respective time of birth. This can be done anonymously through a web interface accessible by scanning a QR code provided in each labor and treatment room or entering the webpage directly. The employee must enter the date, time and room of the capture of the video that is requested deleted and confirm that he or she was participating.

#### Infrastructure for data processing and storage

Figure 4 shows an overview of the data collection infrastructure at SUH for the NewbornTime project.

**Figure 4.**
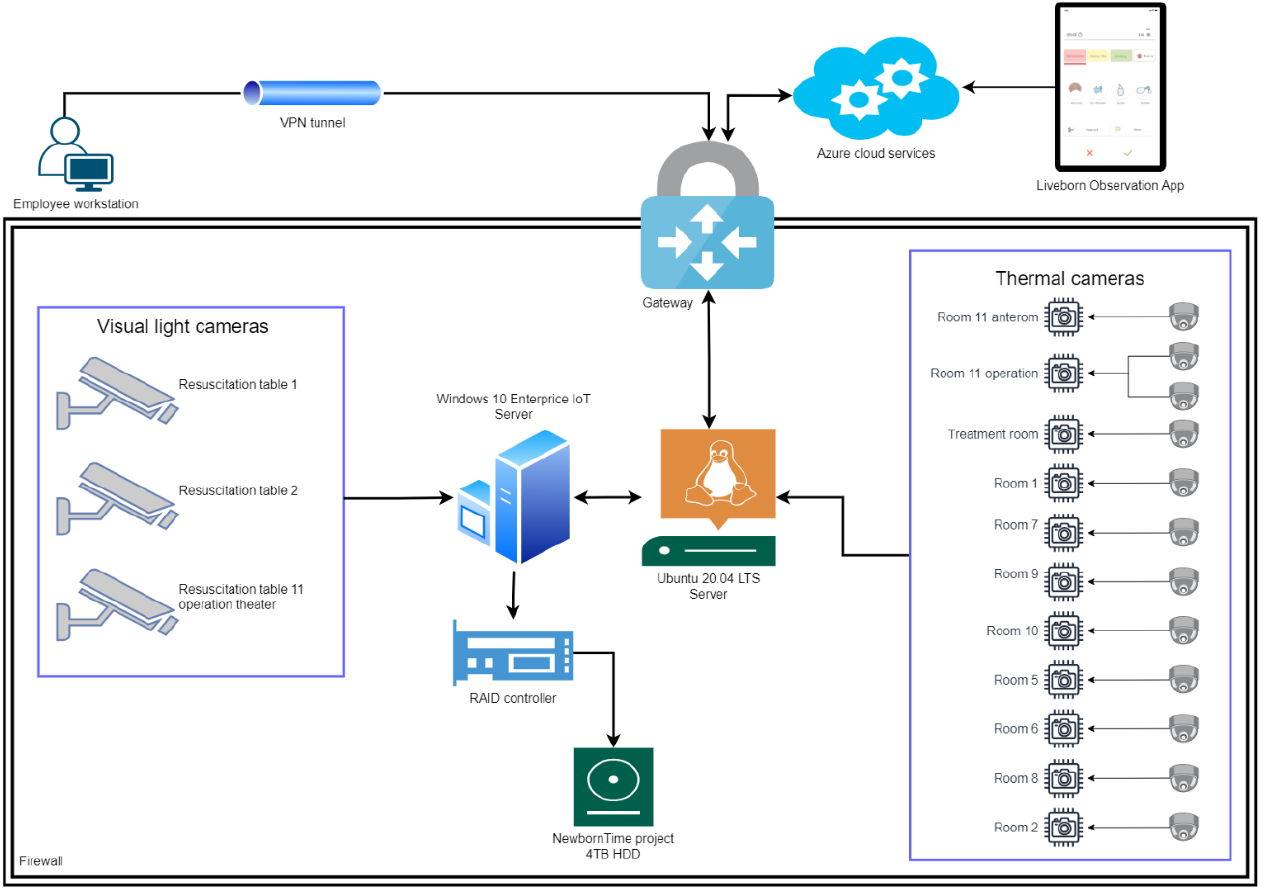
Data collection infrastructure at SUH

The project has 12 thermal cameras and 3 visual light cameras, along with 2 servers. One server is a Windows 10 Enterprise server, while the other is an Ubuntu 20.04 LTS server. The Windows server was set up for an earlier project and is reused now for hosting the data storage unit, which includes a RAID level 5 controller with 8 TB of effective storage, where 4TB are allocated specifically to this project. The Linux server runs the software developed for NewbornTime. All software hosted on this server runs as containerized Docker applications, handling data collection from the thermal cameras as well as data processing and deletion processes. The data captured by the visual light cameras is directly stored on the RAID.

All IoT devices and servers are communicating on a closed network, physically separated from existing IT infrastructure at SUH. This means that access to this network and all resources within it is managed by the project. All communication going in and out of the network is controlled by a gateway where a selected few can access the system via VPN tunnels on a need-to-know basis. Communication with Azure cloud services is also managed by VPN connection, and a DNS server is implemented in the Azure services to resolve domain names.

Internally, the cloud infrastructure consists of multiple virtual networks and subnetworks allocated to different resources. This means that the data storage resources (Database, Blob storage) are segregated from the rest of the infrastructure. Connections between various resources within Azure are established using private links.

Researchers at UiS can access video data stored on the Blob storage through a VPN communication channel established between the blob storage and the local secured server at the university. This data transfer is essential to ensure that the data is accessible for computationally intensive AI algorithm development in a secure environment.

The infrastructure comprises two public-facing web applications: the Liveborn system, which is responsible for collecting metadata information regarding births, and a web application that enables employees to submit opt-out requests.

#### Data flow/processing

Thermal video is captured when the temperature in the measurement area exceeds 30° C. Visual light video is captured while motion is detected. The captured data is stored locally on the RAID storage.

When the Liveborn Observation App uploads data to the Liveborn Web, key data points like ToB, room number and assigned study id are stored in a message queue for every birth. This queue is processed every 6 hours and leads to video data with consent being transferred to cloud storage. The detailed process steps are shown in Figure 5.

**Figure 5.**
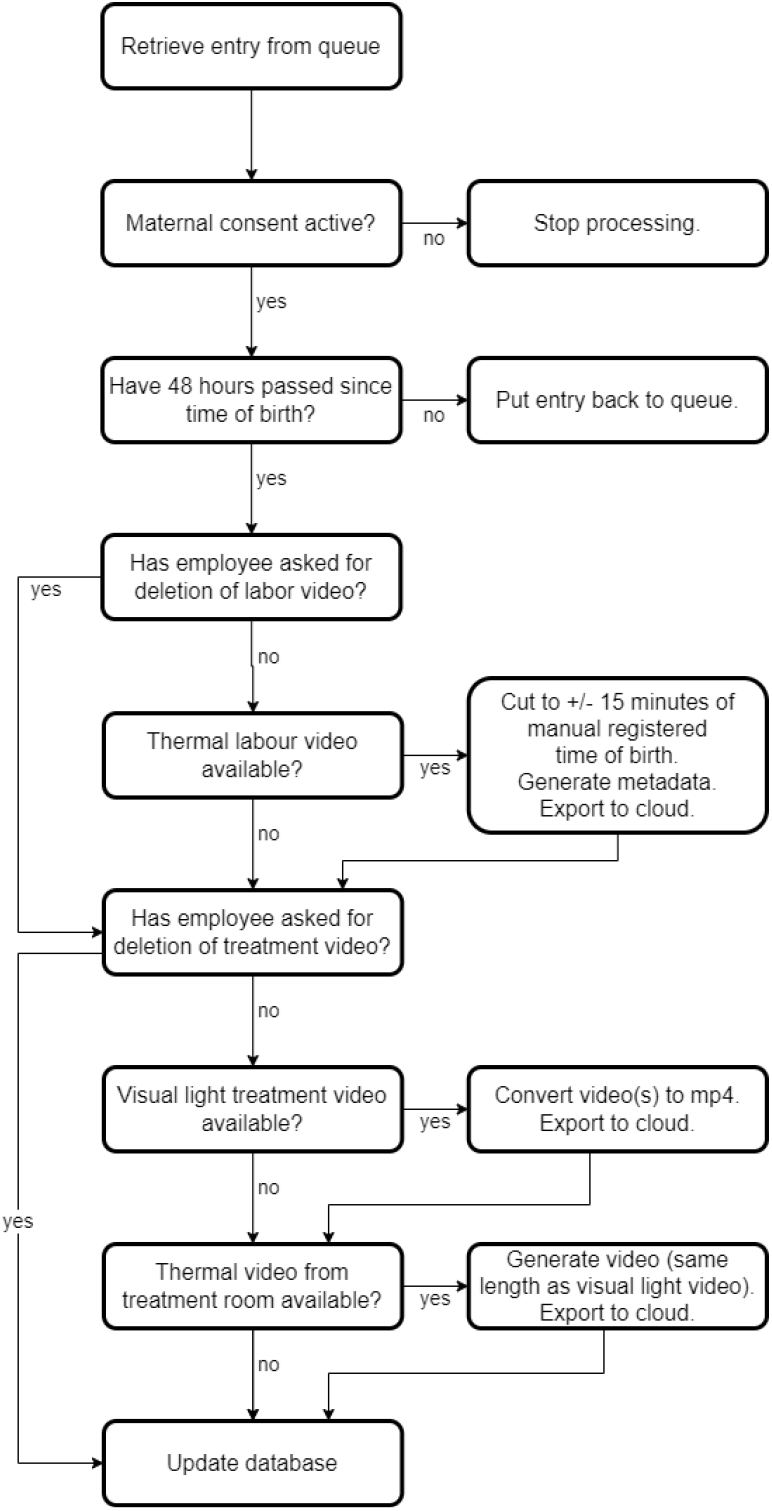
Data processing for video data. The process is repeated every 6 hours.

All video data older than 72 hours is automatically removed from the local storage. If information from the Liveborn App is uploaded late and not reaching the queue processing within 72 hours, this video data will be lost.

If a mother withdraws her consent after data has been collected, the digital consent system will inform the data processor and the data for this mother will be deleted.

#### Amount of data

From November 2021 to July 2024, 2735 women consented to participate of which 2634 extended their consent to allow for educational use of the collected data.

In the same period, 645 thermal videos from birth and 186 visual light videos from resuscitation tables have been collected, with 38 of these videos documenting instances of PPV.

For 157 newborns thermal video from birth and visual light video from treatment at the resuscitation table are available. In 31 of these cases the newborn received PPV, in additional 29 videos the newborn received continuous positive airway pressure (CPAP)

The rate of deletion requests dropped from almost 7.6 /month in the first 6 study months to 0.6 /month in the first 6 months of 2024. Deletion requests from employees were the reason for not processing data in 21 cases.

Data collection is still ongoing with a planned end in Summer 2025.

## Discussion

### Semi-Automated data collection

Video recording newborn stabilization treatment for quality improvement purposes is done in multiple sites around the world, but is still far from scalable to routine practice in most places [15–17]. Continuous video recording and button-triggered recording are commonly used methods for capturing real-life emergency team footage for auditing [2]. Our data collection system incorporates both techniques. It automatically and reliably records thermal and visual light videos. However, a nurse needs to press an on-screen button at the ToB to activate the data processing. Within 72 hours after birth, a research assistant needs to enter additional information on study participation, which triggers automatic verification of consent. If consent is given, transfer of the appropriate video data to the cloud storage. These manual interventions ensure that only data from actual births, with maternal consent, are processed and avoids the need for researchers to review continuous videos that may include mothers and newborns who did not provide consent.

When data collection for NewbornTime was started, there was already an ongoing study called NeoBeat efficacy study [18] that recorded newborn stabilization and resuscitation. To streamline our efforts, we opted to utilize the existing infrastructure and incorporate a digital consent processing and thermal video processing step. The activation of the “Baby born” button at birth was initially introduced by the NeoBeat efficacy study at SUH. We utilize the information this provides to determine the start and end timestamps for cutting the thermal video. Kolstad et al. [19] showed that the button is pressed with a median deviation of less than 2 seconds compared to ToB annotation from manually studying thermal videos.

We experienced data loss when manual inputs were forgotten or not timely entered, especially in holiday seasons with decreased availability of qualified personnel and higher workload of the remaining staff. Once the ToB detection algorithm is developed and implemented, manual interaction at birth will be unnecessary and data collection automated to cover all births with consent.

Currently the only physiological parameter that is collected time synchronously with the videos is newborn heart rate from the NeoBeat heart rate senor device (Laerdal Medical AS, Norway). In future, the Liveborn system could be expanded to include more vital signs like ventilation rates or SpO_2_.

### Consent

In their study, Gelbart et al. [20] examined the ethical and legal aspects of video recording during newborn resuscitation. They highlighted the challenge of obtaining consent during antenatal care, as it could potentially cause unnecessary anxiety for parents. On the other hand, obtaining consent during or after labor may not provide a fully voluntary choice due to the emotional state of the parents at that time.

In the NewbornTime project, the regional ethical committee required antenatal consent from the pregnant women. While the project was enthusiastic about a digital, self-serviced consent solution designed to be user-friendly and secured by blockchain technology, we learned that the pregnant women rarely used the opportunity to sign up proactively although study information was presented in brochures and videos in the waiting areas for antenatal controls. When nurses and research assistants started personally informing women about the ongoing study, the consent rate within that group increased to about 90%, which aligns with consent rates reported for video usage in quality improvement studies [2]. While we experience high percentage of consent for pregnant women who are approached by a project member, the project did not have enough resources to cover personnel costs to ask all expecting mothers for consent.

At the beginning of the project, a significant number of employees requested the deletion of video data, particularly visual light videos captured from the resuscitation table. However, over time, these deletion requests have decreased.

### Better documentation

In their review, Avila-Alvarez et al. [1] concluded that the current quality of documentation in newborn resuscitation is unsatisfactory, highlighting the need for consensus guidelines and innovative solutions. While retrospective documentation may be less accurate than video recordings [2], real-time documentation has shown promising results in capturing interventions during newborn resuscitation [3].

The NewbornTime project aims to provide timelines which would document the ToB, key stabilization/resuscitation activities and newborn heartrate, without the sensitivity of video recordings. These timelines could be used for documentation, input for clinical debriefing and research on guidelines compliance and treatment effectiveness when combined with outcome data.

Video recording newborn resuscitation is standard care in a hospital in the Netherlands where the video is stored in the patient records [15], but the majority of hospitals are only using video recordings for quality improvement initiatives, deleting the videos after some days or weeks [17,21,22]. Integrating videos into the patient journal may raise privacy concerns and resistance among staff. However, using videos exclusively to automatically create precise timelines could effectively address privacy concerns while still preserving crucial information about the patient’s treatment.

### Less concerns on privacy

As thermal videos measure the temperature of objects, it is well suited to recognize the newborn, since it will be the warmest body in the room at birth. Initially we intended to use an absolute temperature measurement threshold to determine ToB but recognized early that too many confounding factors in the delivery room did not allow for collection of data with absolute measurements. These findings are documented by Garcia-Torres et al. [13].

Thermal videos have the benefit of being privacy-preserving for both the patients and the healthcare providers and will minimize identifiable information compared to visual light video. Thermal imaging may be a way forward for labor room recordings to get acceptance from both mothers and hospital staff and comply with privacy regulations.

### Future perspectives

While video review and annotation are time consuming activities, the future AI generated timelines of birth and resuscitation activities do not contain sensitive information and could easily be anonymized and used for research or quality improvement initiatives. Automatic data collection could also allow for scaling data collection and increasing the amount of data that can lead to a better understanding of the time critical activities around birth.

Automated timelines can be incorporated in debriefing systems, quality improvement systems, and retrospective analysis for evidence-based research around newborn resuscitation guidelines and efficiency of treatment.

We believe that the integration of thermal imaging and AI-based activity recognition from video can be extended to other non-diagnostic healthcare applications, such as documenting activities in emergency care or security surveillance, while maintaining privacy for both healthcare providers and patients.

## Conclusion

This work describes the data collection system for recording thermal videos from the labor room and time synchronized visual light videos from newborn stabilization or resuscitation. The system has successfully recorded hundreds of videos from labor and newborn resuscitations; however, the solution could be improved with regards to fully automated consent and data collection process.

## Data Availability

Data used in this research includes videos from labor and newborn resuscitation. Due to the potentially identifiable nature of the dataset the authors are prevented from publicly sharing video data under the ethical approvals for this study. For more information, contact

## Acknowledgment

We express our sincere appreciation to all the women who participated in this study, placing their trust in us and allowing us to utilize their data. A thank you goes out to all the dedicated employees at the maternity ward of SUH for their support.

